# Characterization of Sleep, Spectral, and Epileptiform Activity in Individuals Aged 8–27 Years with SYNGAP1-Related Disorder Using an at-Home Dry EEG Headband

**DOI:** 10.64898/2026.10.21.26366120

**Authors:** Timothy C. Sheehan, Emily L. Schwartz, Kim Paolo Laberinto, Rebecca K. Reh, Maya Dorsey, Katherine McGuckin, Srihari Y. Sritharan, J. Michael Graglia, Virginie McNamar, Kathryn Ann Helde, Albert Misko, Anthony J. Linares, Colin J. Dinsmore, Jake Donoghue, David W. Matthews, Jay Pathmanathan

**Affiliations:** Beacon Biosignals, Boston, MA, USA; CURE SYNGAP1, San Diego, CA, USA; Novartis Institutes for Biomedical Research, Cambridge, MA, USA; BioMarin Pharmaceutical, San Rafael, CA, USA

## Abstract

SYNGAP1-related disorder (SYNGAP1-RD) is associated with sleep disturbances, seizures, and neurophysiological abnormalities. However, in-lab EEG can be difficult to obtain due to the behavioral and sensory disorders associated with this condition. In this study we collected EEG in SYNGAP1-RD participants (n=30 enrolled, 27 contributing data) in a decentralized protocol using a dry electrode headband as a path to longitudinal, research-grade data collection in this population. At-home data collection was successful with 180 recordings containing over 1 hour of usable data collected from 23 participants. Across these recordings, sporadic interictal epileptiform discharges were present in 22 of 23 participants but electrographic seizures were not observed. Relative to an adolescent control population (n=10), SYNGAP1-RD participants showed increased wake after sleep onset, reduced sleep efficiency, and decreased time in N1 and N3 sleep stages along with increased time in N2. We reproduced prior findings from awake spectral EEG analyses, including elevated broadband power along with reduced relative alpha power and alpha-theta ratio in the SYNGAP1 cohort. Notably, differences in spectral power were tightly linked to sleep stage, revealing sleep-stage-dependent spectral biomarker dynamics not previously characterized in SYNGAP1-RD. Finally, we examined differences between variants producing truncations downstream of the GAP domain and those producing truncations before or within it, but analyses were limited by a small sample size (n=4 for the post-GAP variant). Overall, this study demonstrates the feasibility of at-home multi-night sleep EEG collection in participants with SYNGAP1-RD and led to both corroborative and novel findings.

## Introduction

SYNGAP1-related disorder (SYNGAP1-RD) is a Developmental and Epileptic Encephalopathy (DEE) from haploinsufficiency of the SYNGAP1 gene, which encodes a critical regulator of excitatory/inhibitory balance in neuronal signaling and synaptic plasticity. The clinical phenotype is characterized by moderate-to-severe intellectual disability, autism spectrum features, motor impairment, and drug-resistant epilepsy (Graglia et al., 2025; Vlaskamp et al., 2019).

Neurophysiological biomarkers hold considerable promise for tracking disease severity and treatment response in a variety of DEEs, including SYNGAP1-RD. EEG-derived spectral features, such as alpha-theta and alpha-delta power ratios, have emerged as candidate biomarkers. A recent study has shown that quantitative models accurately predicted a SYNGAP1-RD diagnosis (AUC = 0.86) against controls, with the wake alpha-theta ratio in occipital and parietal electrodes among the most informative features (Galer et al., 2025). Beyond resting-state spectral features, SYNGAP1-RD are characterized by disrupted posterior connectivity with abnormal occipital alpha-theta ratios and diminished sleep spindle coherence, suggesting that overnight EEG may yield additional disease-relevant signal not captured during brief awake recordings (Buller-Peralta et al., 2022; Jimenez-Gomez et al., 2019). Sleep-specific spectral features have not been examined in detail in humans, and sleep EEG is also of intrinsic interest given the high prevalence of sleep disturbances in this population: one study found approximately 62% of individuals with SYNGAP1-RD experience sleep disorders (Smith-Hicks et al., 2021). Recently, sleep abnormalities have been identified as potentially characteristic biomarker profiles of neurogenetic disorders, with sleep pathology correlating with severity of psychiatric disease burden (Reyes et al., 2026).

Despite the potential value of both resting wake and sleep EEG, obtaining high-quality neurophysiological data in SYNGAP1-RD is technically challenging. Sensory differences are increasingly reported in SYNGAP1-RD, and managing behavioral outbursts represents one of the most significant sources of stress for caregivers. These behavioral and sensory features frequently preclude standard clinical EEG acquisition, which requires prolonged cooperation with electrode placement and gel application. Severe behavioral problems have been explicitly cited as a technical barrier to accurate EEG testing in this population (Vlaskamp et al., 2019).

Objective sleep characterization has similarly been constrained by these challenges. Actigraphy has been explored as a more tolerable alternative: it is feasible for assessing sleep-wake cycles in children with SYNGAP1-RD, with findings including greater wake after sleep onset, higher fragmentation index relative to typically developing children, and a biphasic sleep pattern with a more prominent secondary sleep period (Doucoure et al., 2026). However, actigraphy does not capture cortical neurophysiology and cannot characterize sleep stage architecture or epileptiform activity. Formal polysomnography (PSG) remains the gold standard for sleep staging but requires in-laboratory overnight studies that are poorly tolerated in this population, limiting access and longitudinal repeatability.

Advances in dry EEG electrode devices have allowed decentralized data collection in clinical trials and patient care. This study utilizes the Waveband headset (Beacon Biosignals Inc.), a dry-electrode EEG headband with four electrode contacts approximately equivalent to F7, F8, O1, O2 in a 10-20 montage that has been cleared by the FDA for home sleep recording. Here we examined the usability of the Waveband in data collection from participants living with SYNGAP1-RD. The Waveband device has previously been shown to have equivalent performance for sleep staging as in-clinic PSG and previously been utilized in an at-home study for participants with Lennox-Gastaut syndrome (LGS) (Pathmanathan et al., 2026; Savietto et al., 2026). Caregivers (all parents of participants with SYNGAP1 variants) were asked to apply and operate the device on their children who have difficulty tolerating standard clinical neurophysiological testing. The goal of this feasibility and usability study of the Waveband device in a cohort of individuals with confirmed SYNGAP1 pathogenic variants along with similarly aged controls was to determine whether longitudinal, research-grade sleep EEG data can be reliably collected in a decentralized manner in this special-needs population.

## Results

### Study Design

This was a Phase 0 device study to test the usability of Beacon’s at-home Waveband EEG monitoring device in individuals with SYNGAP1-RD. This was a fully distributed clinical study, with e-consent, study operations, data aggregation, data processing and analysis performed centrally by Beacon study staff. An initial target enrollment of 30 was sought of participants with SYNGAP1-RD. Participants were instructed to complete up to 10 overnight at-home recordings. This study was collected across two monitoring periods, each comprising 5 consecutive nights of nocturnal sleep monitoring, separated by 2 nights without monitoring for a total of 12 nights (Figure 1A).

**Figure 1.**
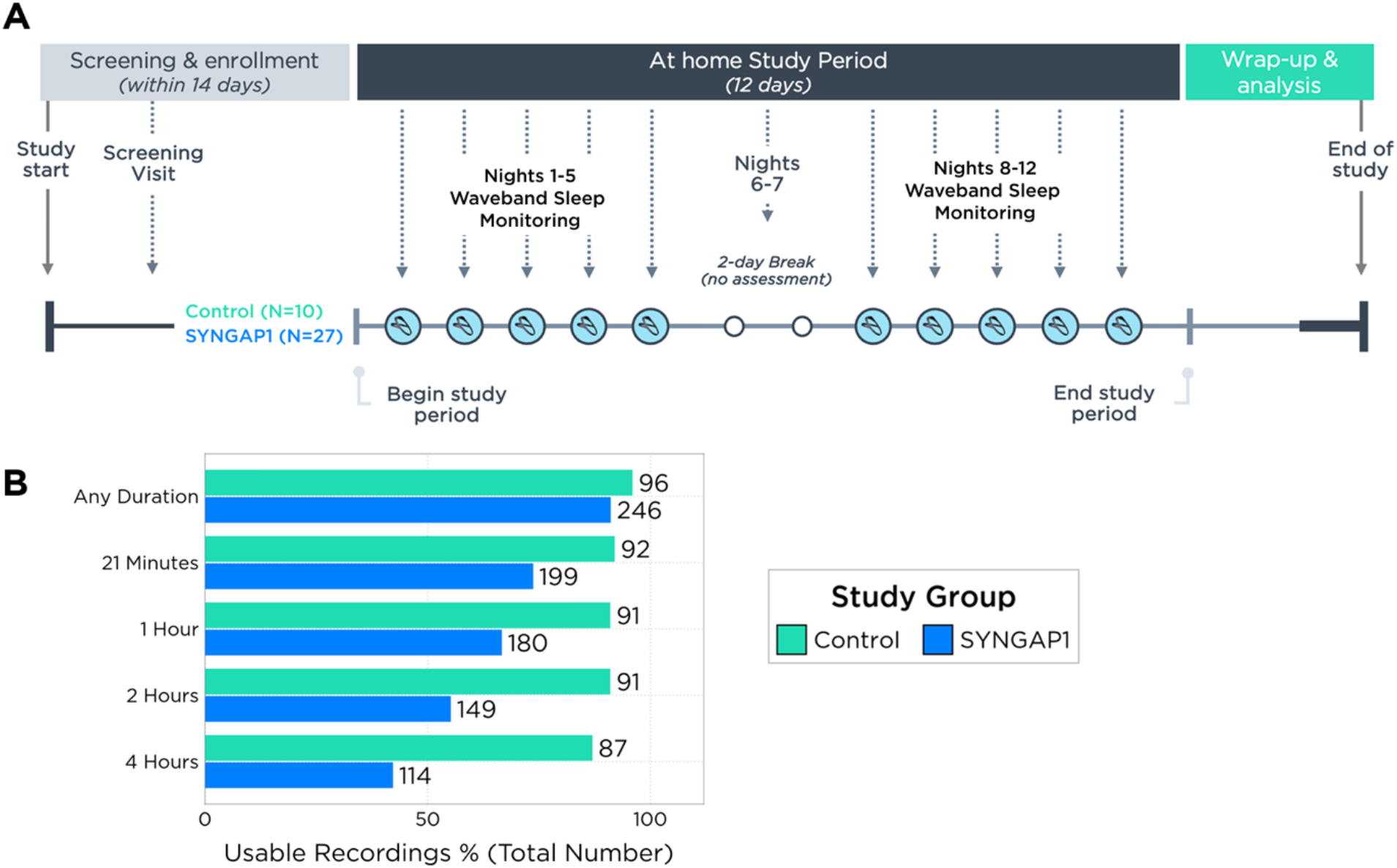
A) Study diagram for at-home overnight waveband recordings. Participants in both cohorts were instructed to complete 10 recordings across a twelve-day period in two blocks separated by a two-day break. B) The percent (bar length) and number (printed value) of all possible recordings completed of different durations across both cohorts along with the number of recordings passing. The control cohort (n=10) has a maximum of 100 completed recordings. The SYNGAP1 cohort (n=27) had a maximum of 270 completed recordings.

### Demographics

Comparative analyses in this study come from two distinct cohorts. The SYNGAP1-RD population was recruited through the CURE SYNGAP1 Foundation specifically for this study targeting a minimum age of 8 years old. The control population consisted of neurotypical siblings of people living with Lennox-Gastaut syndrome that were captured in a previous study (Pathmanathan et al., 2026). Hereafter, the SYNGAP1-RD sample recruited for this study will be referred to as the SYNGAP1 cohort. Both cohorts completed an identical protocol. The population breakdown between SYNGAP1 (n=27) and control (n=10) groups was similar. The SYNGAP1 cohort had a higher proportion of female participants (15/27 vs. 4/10) and a wider age range (8-27 vs. 11-19) but a very similar mean age (14.7 vs. 14.2). Survey data from the SYNGAP1 cohort revealed that 87% (20/23 survey respondents) had a prior epilepsy diagnosis resulting in them taking anti-seizure medications.

### Compliance

The majority of caregivers and parents were able to collect meaningful data from participants living with SYNGAP1-RD. Three of the original thirty target participants were sent headbands but withdrew before initiating any recordings. All other participant families uploaded at least 1 recording, indicating that the Waveband device and Beacon Pal app were usable by the majority of SYNGAP1-RD participants. Sensory and behavioral disorders in this population make EEG data acquisition difficult. Nevertheless, 23/27 participants (85%) who uploaded any data were able to upload at least a single recording with over one hour of usable data (180 recordings total, Figure 1B). This threshold corresponds to an extended clinical EEG as well as inclusion in our analyses of spectral activity, sleep onset characteristics (latency to persistent sleep and sleep onset latency), and the presence of epileptiform features. Note that 9/180 recordings that had >1 hour of usable data were ultimately excluded from the EEG BIOMARKER set due to having a high percentage of poor quality or off-head data in the middle of the recordings, leading to 171 recordings (from 23 participants) included in the EEG BIOMARKER set (see Methods). Lastly, 20/27 participants (74%) completed at least one recording with over four hours of usable data (114 recordings total, Figure 1B) meeting the minimum AASM criteria for an overnight PSG. Ultimately 79 of these recordings (from 19 participants) were included in our SLEEP BIOMARKER set after excluding recordings with high uncertainty in TST or WASO (see Methods). These recordings were included in our analysis of sleep endpoints. The control cohort uploaded a total of 96 recordings of which 90 were included in the EEG BIOMARKER and 78 were included in the SLEEP BIOMARKER datasets. The higher compliance from the control cohort points to challenges associated with collecting data from the SYNGAP1-RD population while the high number of usable recordings collected overall points to the utility of the at-home repeated night paradigm.

### Sleep

Aggregate sleep data across control and SYNGAP1 recordings meeting our inclusion criteria showed some notable differences. While participants in both cohorts slept for a similar total amount of time (TST, Figure 2A), SYNGAP1 participants showed altered sleep architecture, with sleep efficiency (SE) being significantly lower (by 5.21%, FDR-corrected p < .05), and had notable differences across sleep stages. Specifically, relative to the control cohort, the SYNGAP1 group spent less time in N1 and N3 (by 1.51% and 5.33%, respectively; both FDR-corrected p < .05) and more time in N2 (by 8.53%; uncorrected p < .05), with the reduction in deep sleep (N3) being of particular interest (Figure 2B). No significant differences were found between the groups for the percent of time spent in REM (1.55% lower in SYNGAP1 group, uncorrected p > .05), indicating sleep stage disruption was specific to NREM. Interestingly, SYNGAP1 participants seemed to enter persistent sleep 6.8 minutes earlier suggesting no difficulty initiating sleep at the start of the night (Figure 2C, uncorrected p > .05). However, wake after sleep onset (WASO) was significantly greater in SYNGAP1 participants with an increase of 17.3 minutes compared to controls (FDR-corrected p < .05). Sleep transition structure was largely preserved between groups, with only negligible differences in the sleep fragmentation index (SFI, 1.35 lower in SYNGAP1, uncorrected p > .05) and awakening index (AWI, 0.3 higher in SYNGAP1, uncorrected p > .05; see Figure 2D). The significant increase in WASO aligns with previous reports of more disrupted sleep in SYNGAP1-RD (Doucoure et al., 2026).

**Figure 2.**
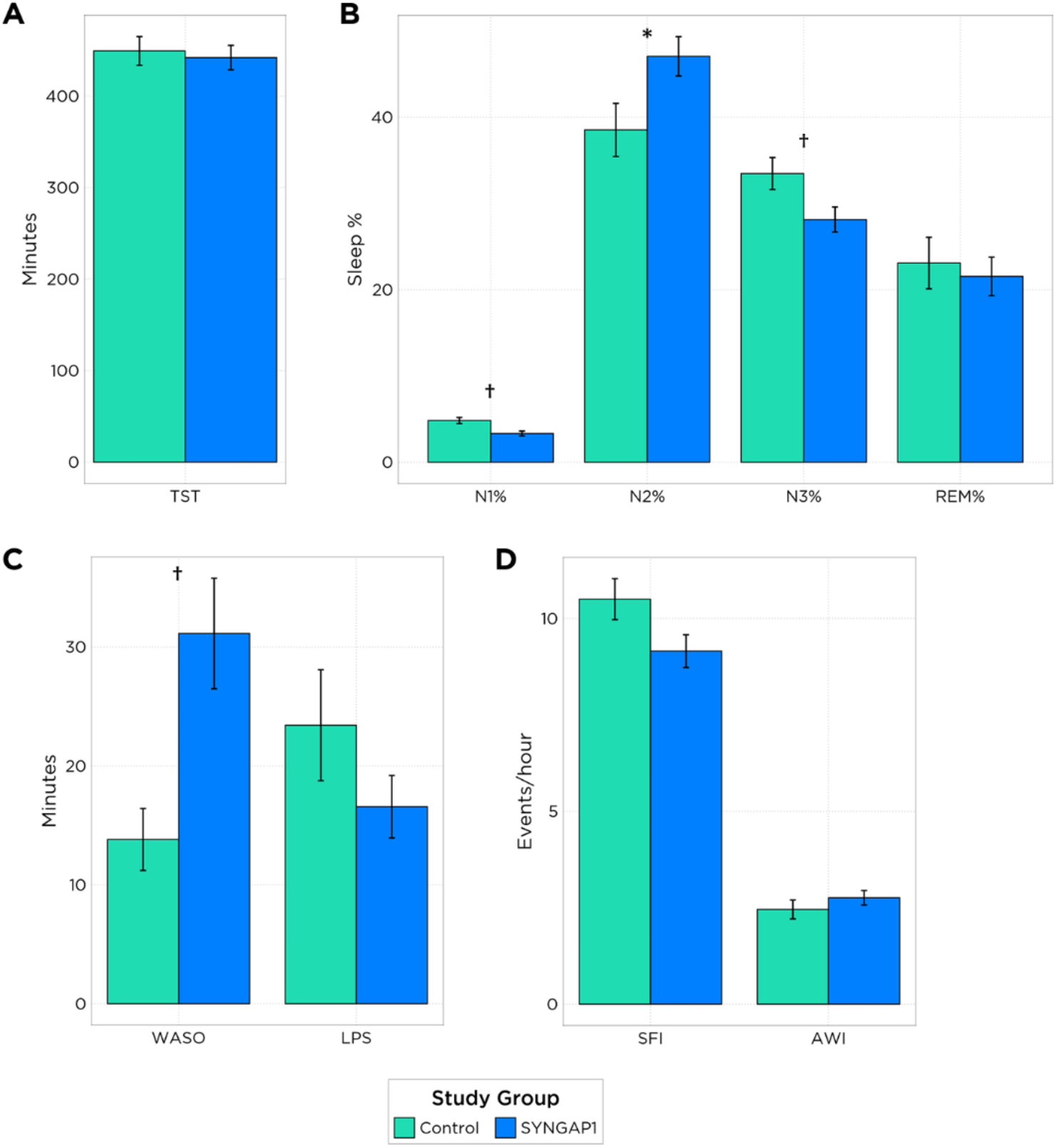
Overview of sleep macroarchitecture between groups. Bar heights reflect model estimated mean (while accounting for age and interaction of age and study group) and error bars reflect model estimated standard errors. A) Total sleep time (TST) in minutes per group. B) Percentage of sleep time spent in each stage per group, with SYNGAP1 participants spending significantly less time in N1 and N3. C) Wake after sleep onset (WASO) and latency to persistent sleep (LPS) in minutes per group, with SYNGAP1 participants showing significantly greater WASO. D) Sleep fragmentation index (SFI) and awakening index (AWI) in events/hr per group. Significant group differences indicated by † FDR-corrected p < 0.05; * uncorrected p < 0.05. Visualized metrics are derived from the SLEEP BIOMARKER dataset (n=157 recordings), with the exception of LPS which was derived from the EEG BIOMARKER dataset (n=261 recordings, see Methods).

### Spectral Analyses

Recent work evaluated differences in spectral activity in SYNGAP1 individuals and identified both alpha-theta and alpha-delta ratios as highly diagnostic of the disorder (Galer et al., 2025). To first align with this approach, which utilized 10-20 EEG collected in awake in-clinic recordings, we used the wake portion of the overnight recordings. We replicated findings with reduced alpha-theta and alpha-delta ratios during wake in the SYNGAP1 cohort (p<.05, uncorrected, Figure 3F, Table 2), confirming this prior work in an at-home setting using the Waveband. In addition, we found reduced relative alpha and increased broadband power in the SYNGAP1 cohort (uncorrected p < .05) but no significant differences in delta, theta, or beta power (uncorrected p > .05).

**Table 1:** Model based estimates of sleep features across study groups while accounting for age. Values depict Mean (SEM) for each group or the difference between groups. * p < 0.05 (uncorrected); † p < 0.05 (FDR-corrected)

| Feature | Control (n=10) | SYNGAP1 (n=19) | Difference | Sig. (p) |
| --- | --- | --- | --- | --- |
| WASO | 13.81 (2.6) | 31.13 (4.65) | 17.32 (5.33) | † |
| TST | 449.42 (15.65) | 442.14 (13.36) | -7.27 (20.58) |  |
| SFI | 10.5 (0.53) | 9.15 (0.42) | -1.35 (0.68) |  |
| SE | 90.61 (1.51) | 85.4 (1.28) | -5.21 (1.98) | † |
| REM% | 23.09 (2.99) | 21.55 (2.23) | -1.55 (3.73) |  |
| N3% | 33.47 (1.85) | 28.14 (1.45) | -5.33 (2.35) | † |
| N2% | 38.53 (3.08) | 47.06 (2.28) | 8.53 (3.83) | * |
| N1% | 4.84 (0.36) | 3.33 (0.28) | -1.51 (0.45) | † |
| LPS | 23.42 (4.66) | 16.57 (2.63) | -6.85 (5.35) |  |
| AWI | 2.46 (0.25) | 2.76 (0.19) | 0.3 (0.31) |  |

**Table 2:** Model based estimate of having SYNGAP1-RD on wake spectral power while accounting for age. Values reflect relative power for individual spectral bands or spectral power ratios. * p < 0.05 (uncorrected); † p < 0.05 (FDR-corrected).

| Spectral Feature | Effect size (z) | Sig. (p) |
| --- | --- | --- |
| Delta | 1.91 |  |
| Theta | -1.91 |  |
| Alpha | -2.11 | * |
| Beta | -1.39 |  |
| Broadband | 2.26 | * |
| Alpha-theta ratio | -2.02 | * |
| Alpha-delta ratio | -2.08 | * |

**Figure 3.**
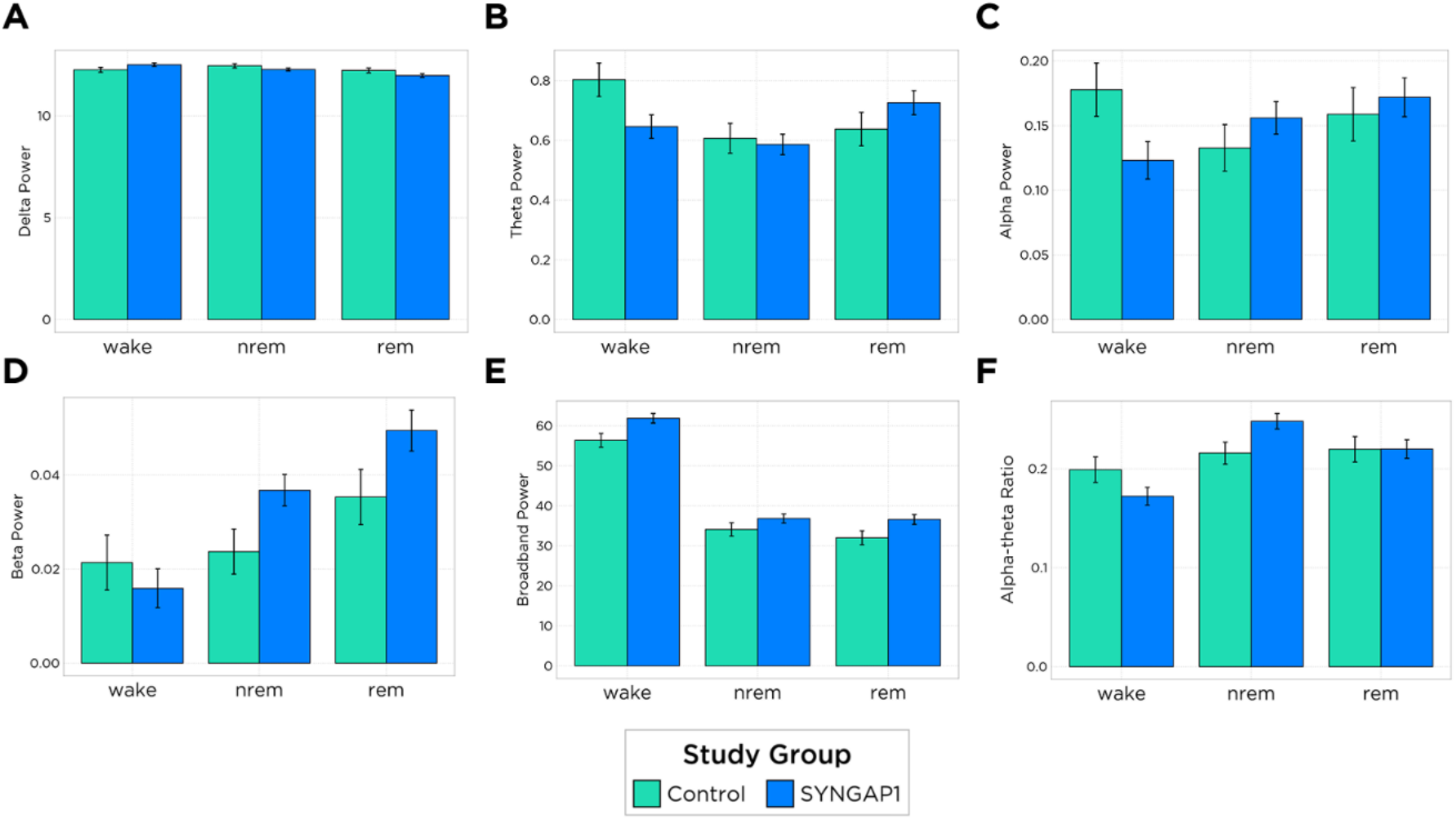
Sleep stage interacts with spectral differences. Displayed values reflect model estimated mean and standard error. Power is shown across wake, NREM, and REM sleep stages for each group. Relative power for A) delta, B) theta, C) alpha, D) beta, along with E) broadband, and F) alpha-theta ratio.

The overnight recordings captured here have additional information over in-clinic daytime routine EEG as they capture spectral activity across sleep stages. To evaluate if the pattern of spectral activity during sleep is different between SYNGAP1 participants and controls, we compared two models using a likelihood ratio test. A complete model including the interaction of stage and population was compared with a simpler model including stage but not the interaction term. This approach across spectral bands revealed a significant improvement in model fit for delta, theta, alpha, beta, broadband and alpha-theta ratio (p < .05, FDR-corrected; Table 3, Figure 3). Notably alpha-theta showed by far the largest fit improvement when measured by both the likelihood ratio test and change in Bayesian information criterion (BIC) suggesting alpha-theta ratio is particularly sensitive as a biomarker when measured across sleep stages.

**Table 3:** Likelihood-ratio test model for the impact of SYNGAP1 diagnosis interacting with sleep stage. P-values correspond to χ^2^ comparing model fit with 2 degrees of freedom. † p < 0.05 (FDR-corrected).

| Spectral Feature | $\chi^2$ | ΔBIC | Sig. (p) |
| --- | --- | --- | --- |
| Delta | 20.24 | -3.21 | † |
| Theta | 19.16 | -2.13 | † |
| Alpha | 19.96 | -2.93 | † |
| Beta | 10.65 | 6.38 | † |
| Broadband | 10.87 | 6.16 | † |
| Alpha-theta ratio | 29.87 | -12.84 | † |
| Alpha-delta ratio | 4.35 | 12.69 |  |

### Epilepsy

Epileptiform activity was limited across the 171 recordings in the EEG BIOMARKER dataset. Sporadic interictal epileptiform discharges (SIED) were the most frequently observed epileptiform pattern, identified in 22 of the 23 participants (95.7%) with analyzable recordings (Figure 4A). A trend toward decreasing SIED frequency with age (−0.13 (events/minute) per year of age) was observed but did not reach statistical significance (p = 0.12, Figure 4B). No seizures or lateralized periodic discharges (LPDs) were observed across the entire cohort. Generalized paroxysmal fast activity (GPFA) was observed in one participant (1/23, 4.3%). Additionally, sharp wave discharges (SWD) were similarly rare, identified in one participant (1/23, 4.3%), and generalized periodic discharges (GPDs) were observed in two participants (2/23, 8.7%). Notably, the four participants demonstrating non-IED epileptiform patterns (GPFA, SWD, GPD) were among the younger individuals in the SYNGAP1 cohort (ages 8, 9, 12, and 12 years). This may suggest epileptiform activity decreases with age or alternatively that older participant’s epilepsy is better controlled with anti-seizure medications.

**Figure 4.**
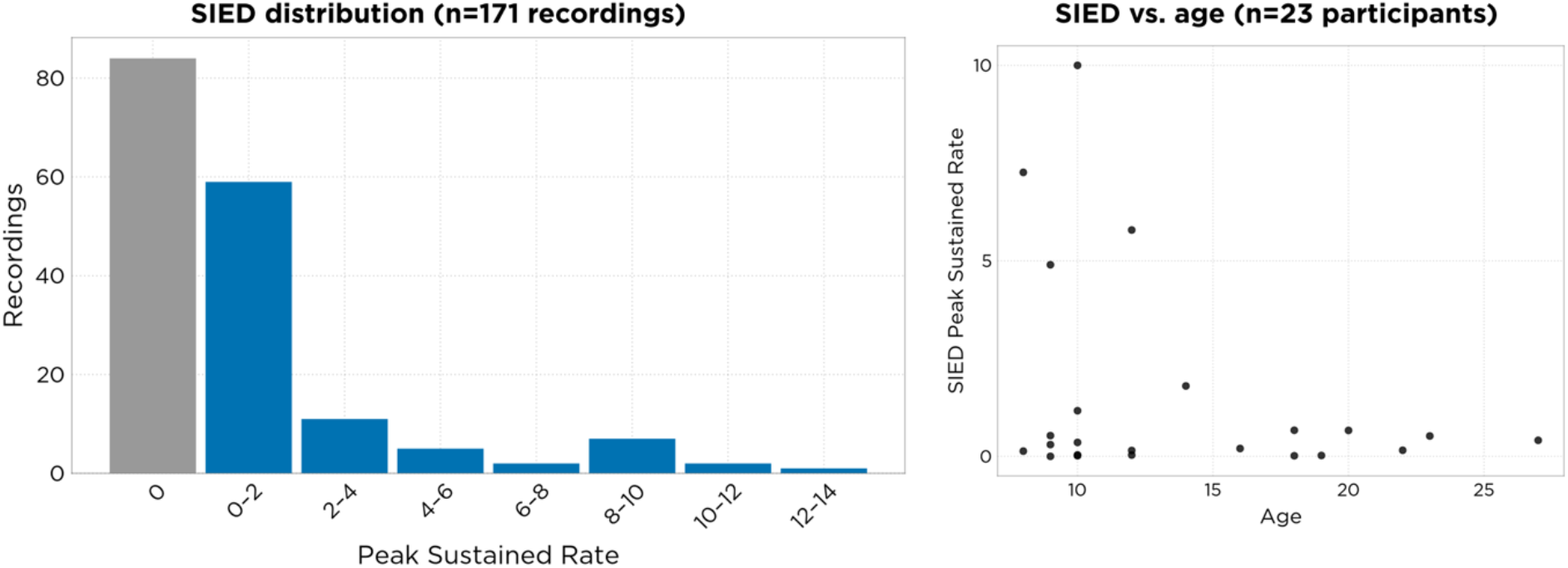
A. Peak sustained rate of SIEDs as reported by recording level overreads by a human expert (with Neurologist oversight). B. Scatter plot of mean peak rate of SIEDs across recordings for each participant vs. age.

### Genetics Results

The evidence for genotype-phenotype correlations in SYNGAP1-RD is limited. Vlaskamp et al. (2019) reported that, across their cohort and previously published cases, milder intellectual disability was more common with variants in exons 1-4 than in exons 5-19. Intellectual disability severity did not differ significantly by variant type, and clinical features were described as broadly similar across truncating, splice-site, and missense variants. However, this study relied on clinical outcome measures, and whether variant location relates to objective neurophysiological features such as sleep EEG has not been examined. As an exploratory analysis, sleep architecture and spectral EEG features that showed significant differences in the full SYNGAP1 cohort were examined across two variant location subgroups to assess whether findings varied by variant location. In the SLEEP BIOMARKER set and of the 17 participants that had genetic reports, 15 carried protein-truncating variants and 2 carried missense variants. Participants carrying missense variants were assigned to the pre/within-GAP subgroup based on residue position. Of the 19 participants with genetic reports in the SPECTRAL BIOMARKER set, 17 carried protein-truncating variants and 2 carried missense variants. Spectral features were selected based on features of interest from the previous literature (Galer et al., 2025) as well as broadband power. Sleep architecture comparisons included n=14 pre/within-GAP and n=3 post-GAP participants; spectral comparisons included n=15 and n=4, respectively. While no effects were significant following FDR-correction, REM% was lower for the post-GAP group relative to the pre/within GAP group (uncorrected p < .05) and was the only sleep or spectral feature to show any uncorrected significant differences between genetic subgroups (see Table 4, Supplementary Figure 1). Recording yield and quality did not differ significantly between genetic subgroups, including number of usable recordings, percentage of usable recordings, and total recording duration (uncorrected p > .05).

**Table 4.** Sleep architecture and spectral EEG features across controls, pre-/within-GAP, and post-GAP variant subgroups. Sleep architecture comparisons: control n=10, pre-/within-GAP n=14, post-GAP n=3. Spectral comparisons: control = 10, pre-/within-GAP n=15, post-GAP n=4. For each feature, the median (25th–75th percentile) is reported per group. Between-group differences were assessed with two-sided Mann-Whitney U tests for the control vs. pre/within-GAP, control vs. post-GAP, and pre/within vs. post-GAP comparisons; effect sizes are reported as the rank-biserial correlation (r), where a positive r indicates higher values in the first-named group of each comparison and a negative r indicates higher values in the second. * p < 0.05 (uncorrected), † p < 0.05 (FDR-corrected).

| Feature | Control | Pre/Within-GAP | Post-GAP | Control vs. Pre/Within-GAP | Control vs. Post-GAP | Pre/Within-GAP vs Post-GAP |
| --- | --- | --- | --- | --- | --- | --- |
| WASO | 12.69 (10.48-22.66) | 43.0 (23.33-60.1) | 49.5 (40.08-69.5) | -0.65 * | -1.0 * | -0.19 |
| SE | 91.12 (88.58-92.83) | 85.13 (82.71-90.67) | 88.55 (80.65-89.03) | 0.44 | 0.53 | 0.14 |
| N3% | 33.54 (30.5-38.12) | 31.16 (20.0-36.9) | 24.0 (21.77-34.27) | 0.33 | 0.4 | -0.05 |
| N2% | 39.68 (36.52-40.7) | 43.79 (36.29-50.76) | 57.81 (47.44-64.09) | -0.29 | -0.6 | -0.48 |
| N1% | 4.34 (3.90-4.85) | 3.30 (2.39-3.75) | 3.60 (3.3-3.62) | 0.59 * | 0.73 | -0.1 |
| REM% | 23.01 (19.87-24.65) | 23.82 (15.98-26.42) | 14.59 (10.52-14.99) | 0.07 | 1* | 0.86* |
| Broadband (wake) | 60.97 (59.64-62.14) | 63.96 (60.26-67.58) | 59.84 (58.27-60.8) | -0.41 | 0.15 | 0.53 |
| Alpha-Theta Ratio (wake) | 0.20 (0.17-0.20) | 0.18 (0.15-0.20) | 0.21 (0.19-0.23) | 0.36 | -0.1 | -0.57 |
| Alpha-Delta Ratio (wake) | 0.02 (0.01-0.02) | 0.01 (0.01-0.02) | 0.01 (0.01-0.02) | 0.44 | 0.1 | -0.4 |

## Discussion

This study demonstrates that longitudinal, research-grade sleep EEG can be collected at home from people living with SYNGAP1-RD (particularly focused on adolescents) using a caregiver-administered, dry EEG electrode headband. This population could benefit from such technology because of the challenges of standard in-lab clinical EEG within the context of sensory and behavioral comorbidities, the value of longitudinal monitoring, and the potential for EEG biomarkers to serve as outcome measures for upcoming disease-modifying therapies currently in consideration. Across 171 recordings from 23 SYNGAP1-RD participants and 90 recordings from 10 control participants, the Waveband device captured sleep architecture, spectral activity, and epileptiform events with sufficient quality to detect group differences from controls and replicated previously reported wake spectral findings, supporting decentralized EEG as a viable approach for biomarker development in this hard-to-study population.

We found SYNGAP1 participants showed reduced sleep efficiency, less N1 and N3, more N2, and greater WASO relative to neurotypical controls, with sleep stage differences confined to NREM. These findings are broadly consistent with prior reports of caregiver-reported disrupted sleep in SYNGAP1-RD, including reduced sleep time relative to controls, and extend this literature by characterizing EEG-based sleep architecture. Notably, we found that NREM-specific stage redistribution was more prominent than global or REM sleep loss, in contrast to our prior work examining sleep in participants living with Lennox-Gastaut Syndrome (Pathmanathan et al., 2026). We additionally found a left shift of spectral frequency content (increased delta power relative to theta, alpha and beta power) in the awake EEG, which has been previously reported and noted as a key feature in EEG-based DEE classifier models (Galer et al., 2025). However, we found this to be a state-specific shift, with the reverse occurring in NREM sleep (EEG power spectra shifted right, with more relative alpha/beta power, see Figure 3). Increased low frequency power is commonly seen across DEEs in the wake state, but we are unaware of prior data demonstrating a relative reversal of this trend in NREM.

One candidate network that may contribute to the elevated broadband and delta power observed during wake, as well as sleep dysregulation and the shift in power toward higher frequency bands observed during sleep, is the thalamic reticular nucleus (TRN). Impaired TRN-mediated sensory gating is a proposed circuit-level endophenotype across neurodevelopmental disorders (Krol et al., 2018), and could plausibly account for increased cortical excitability, altered low-frequency thalamocortical dynamics during wake, and fragmented sleep. Consistent with this, reduced thalamic inhibition in autism spectrum disorder has been linked to both impaired sensory gating and sleep spindle deficits (Baran et al., 2023). Elevated wake delta/broadband power has been reported in other neurodevelopmental populations with presumed thalamocortical involvement, such as autism spectrum disorder (Cornew et al., 2012), though this finding has not been consistently replicated. More recent work has linked sensory processing deficits in SYNGAP1-RD to excitation/inhibition (E/I) imbalance (Zhang et al., 2026). This hypothesis could be further tested by examining EEG sleep spindle characteristics, which are generated by TRN circuitry and should be reduced in TRN dysfunction. This is a natural target for future work using this dataset or platform.

Additionally, epileptiform activity was nearly ubiquitous (with sporadic IEDs in 22 of 23 participants) but was sparse in absolute terms, and there were no seizures identified despite the long recording periods. The nominal decrease in sporadic IED frequency with age observed here contrasts with most published series in SYNGAP1-RD and other DEEs, which typically report IED burden increasing with age. For example Ribeiro-Constante et al. (2024) reported increasing disorganized background activity and generalized discharges with age. This discrepancy could reflect a cohort effect rather than a true departure from the natural history of the disorder: published series generally enroll from infancy, whereas the youngest participants here were 8 years old. A similar attenuation of IEDs with age, without full resolution of the underlying epileptiform tendency, has been described in other DEEs such as Lennox-Gastaut syndrome. Some age-related shifts described in the pediatric epilepsy literature, such as an occipital-to-frontal shift in IED topography, are difficult to evaluate with the frontal temporal/occipital bipolar montage used here and are noted only as context (Kim et al., 2024). The sparse electrode coverage of the Waveband device could miss focal IEDs but is unlikely to miss generalized, frontotemporal, or occipital discharges. Additionally, generalized paroxysmal fast activity was not commonly seen, in contrast to prior reports, but this feature was observed in our prior evaluation of Lennox-Gastaut syndrome (Pathmanathan et al., 2026). If epileptiform activity does attenuate with age in this population, its utility as a stand-alone EEG biomarker may be limited in older cohorts, increasing the relative value of the sleep architecture and spectral measures described above, which showed clearer group separation in this age range. This would need to be confirmed with standard 10-20 clinical EEG.

This study has several limitations. The genetic subgroup analyses were severely underpowered (n=3-4 post-GAP participants) and should be interpreted as hypothesis-generating rather than definitive. The absence of significant differences between pre/within-GAP and post-GAP groups does not rule out a true effect of variant location. Additionally, because location-based grouping is most interpretable for truncating variants, the inclusion of two missense variants in the subgroups may further limit interpretation. The control and SYNGAP1 groups were also unequal in size (n=10 vs. n=23), and caregiver-administered at-home recording, while central to the feasibility aims of the study, introduces variability in electrode placement, necessitates simplified technology that offers sparse electrode coverage, and inherently will be conducted under uncontrolled recording conditions that would not be an issue in the clinical setting. Finally, the cross-sectional age range sampled here limits conclusions about developmental trajectories of epileptiform activity, EEG spectral maturation, or sleep architecture in SYNGAP1-RD.

The amount of data collected shows that the Waveband device offers sensory tolerability, broad accessibility with home Wi-Fi for data upload, and ease of use for caregivers. Overall, the results support the feasibility of decentralized, multi-night EEG in SYNGAP1-RD and identify NREM sleep architecture and sleep-stage-dependent spectral measures, particularly the alpha-theta ratio, as candidate biomarkers warranting further validation in larger, longitudinal cohorts. For appropriate questions, our data indicate that caregiver-collected, limited-montage EEG can provide valuable biomarkers of disease and may be valuable as disease progression and pharmacodynamic response biomarkers as well.

## Methods

### Waveband system

The Waveband (formerly Dreem 3S) is an FDA 510(k) cleared medical device intended for prescription use to measure, record, display, transmit and analyze the electrical activity of the brain to assess sleep in a home or healthcare environment. The Waveband headband and Beacon Pal mobile app work together to provide summary sleep metrics and a hypnogram output of sleep scoring for 30-second recording epochs. These metrics are used to generate predefined reports using participant data to assist in the assessment of sleep in adult individuals aged 22 to 65 years old. While the device has not been cleared for use in pediatric populations, suitable quality recordings are generally possible from this population by additionally placing a fit-band over the headband to maintain good electrode contact and stability on small head sizes.

The Waveband records EEG at a sampling rate of 250 Hz from four electrodes (F7, F8, O1, O2), yielding five bipolar channels: F7-O1, F7-O2, F8-O1, F8-O2, F8-F7. In addition to the five EEG channels, the Waveband also includes a single lead assessment of cardio activity (SLACA), yielding two additional physiological channels: SLACA-F8, SLACA-F7.

### Study compliance

Recordings were scheduled to be completed across two contiguous 5-night blocks separated by a 2-night break. However, scheduling conflicts or technical issues sometimes required recordings to be rescheduled. Rescheduled recordings were assigned to their closest respective study day, with a maximum of 10 recording nights analyzed per participant. Extra recordings, or recordings that otherwise violated the study protocol, were flagged as non-compliant and excluded from subsequent analysis. When a participant recorded more than 10 compliant recordings, only the first 10 were analyzed.

### On-head cropping

All recordings underwent automated cropping, in which periods identified as off-head were removed from the end of the recording. Epochs labeled as off-head represent periods when no electrodes were in contact with the scalp. This label did not necessarily mean the headset had come off the participant’s head entirely, and it was possible for the headset to shift between off-head and on-head status throughout the night without intentional manipulation. This cropping procedure removes the typically brief period where a participant removes the headband in the morning prior to formally stopping the recording in the Beacon Pal app. All cropped recordings underwent manual adjudication to confirm the accuracy of the cropping window.

### Quality control steps

To be included in the SLEEP BIOMARKER analysis set used for all sleep-based endpoints, recordings were required to pass the manual check for protocol compliance, contain at least 2 hours of total sleep time before removal of unusable epochs, contain at least 4 hours of usable time after removal of unusable epochs, have at least 25% of the recording labeled on-head, and show an estimated error of TST and WASO not exceeding 60 and 30 minutes, respectively, as determined by a bootstrapped analysis in which unusable epochs were replaced with random stages.

To be included in the EEG BIOMARKER analysis set used for all spectral, epilepsy, and sleep-onset– based endpoints, recordings were required to pass the manual check for protocol compliance, contain at least 1 hour of usable time after removal of unusable epochs, and have at least 25% of the recording labeled on-head.

### Usable epochs

#### Sleep staging

Inclusion for sleep staging was based on the output of our quality algorithms. A single 30-second epoch was considered usable for sleep staging if it was classified as on-head with at least 1 channel containing 18 or more seconds of good-quality EEG (as determined by a quality algorithm which judges quality based on suitability for staging), or if it was classified as off-head with a Wake probability greater than 75%.

#### Spectral analyses

All epochs labeled as off-head were excluded to minimize the potential impact of artifacts. Because spectral features are sensitive to artifacts related to movement, blinks, and epilepsy-related activity, the EEG was divided into 2-second segments, and segments were marked unusable if root mean square (RMS) signal amplitude exceeded 710 mV, if peak-to-peak RMS amplitude difference exceeded 505 mV, or if signal variance fell below 1 μV^2^. Note these thresholds are three orders of magnitude higher than thresholds used in adult populations and were based on exceeding 5 standard deviations of the sample mean.

After artifact rejection, absolute spectral power was computed over each 2-second segment. Values were then aggregated within sleep stages by averaging across non-rejected segments. Results are reported both qualitatively and through mixed-effects models. A given sleep stage was included in spectral analyses only if it retained at least 90 seconds of usable data following artifact rejection.

#### Frequency bands

The following frequency ranges were used for this analysis: delta (0.4–4 Hz), theta (4–8 Hz), alpha (8–12 Hz), beta (12–30 Hz), and broadband (0.4–50 Hz). Note that gamma (30-50 Hz) was initially computed but was deemed to unreliable to report resulting in the wider broadband range. All ranges are inclusive of the lower band and exclusive of the upper band.

#### Definition of band power

Absolute power in a frequency band was defined as the weighted average of the estimated power spectral density (PSD) over that band’s frequency range, with each frequency bin weighted according to bandwidth. Because absolute broadband power reflects this same weighted average computed across the full frequency range rather than a sum across bands, it could be smaller than the absolute power in any individual sub-band. Relative power in a frequency band was defined as the ratio of the absolute power in that band to the absolute broadband power.

#### Computation of spectral power

Two ipsilateral bipolar EEG channels (F7–O1 and F8–O2) were derived from four electrodes (F7, F8, O1, O2; see Waveband system section). Prior to spectral feature extraction, a fourth-order zero-phase Butterworth 0.3 Hz high-pass filter was applied to minimize the influence of low-frequency drift. The spectral content of the EEG was computed as a spectrogram — a time-resolved estimate of the PSD — by dividing the EEG into consecutive 2-second segments overlapping by 1 second and estimating the PSD for each channel within each segment using a multitaper approach. Segments were rejected according to the artifact rejection criteria described above (see Usable Epochs: Spectral Analyses) prior to computing absolute power. If a segment was rejected for any channel, it was excluded across all channels. Absolute power in each band was then computed for each channel and segment among the remaining, non-rejected segments.

For each channel, the mean value across all non-rejected segments within a sleep stage was computed, yielding one absolute power value per recording, sleep stage, band, and channel. Broadband power was computed as the weighted average across all spectral bands for each channel and sleep stage, accounting for each band’s relative bandwidth. Relative power was computed by dividing each absolute power value by the broadband power value for the corresponding channel and sleep stage. The alpha-theta ratio was computed by dividing mean alpha power by mean theta power for a given channel and sleep stage. The alpha-delta ratio was computed using the same approach. Relative power values were expressed as ratios, while absolute power values, including broadband power, were converted to decibel units (dB = 10 × log10[power in µV^2^/Hz]).

#### Modeling

To identify candidate EEG biomarkers, sleep architecture and spectral measures were compared between SYNGAP1 participants and healthy controls. Effect sizes and significance were evaluated using mixed-effects models, with the feature of interest as the outcome and fixed effects for population (control vs. SYNGAP1), mean-centered age, and the interaction between age and population. A random intercept for each participant was included as the sole random effect, to account for participant-specific attributes underlying repeated measures across days. Unless otherwise noted, reported p-values correspond to the direct effect of SYNGAP1 status. Significance was assessed using the FDR-corrected p-value, calculated separately within each of three feature sets: sleep, wake spectral, and sleep-spectral interactions. For values that were not significant after FDR correction, the nominal p-value was noted instead (p < .05). To evaluate whether the pattern of sleep-stage-related activity differed between SYNGAP1 participants and controls, a separate set of likelihood ratio tests was performed, comparing two nested models: a full model including the interaction between sleep stage and population, and a reduced model including sleep stage but omitting the interaction term. These tests were conducted independently of the main models described above, which evaluated only the direct effect of SYNGAP1 status. For epilepsy features, which were evaluated only in SYNGAP1 participants, a simpler model was fit including only the fixed effect of age, along with a random intercept for participant. All modeling was performed using the MixedModels.jl package (v5.4.0) in Julia (v1.12.0).

### Epilepsy Annotations

All recordings in the EEG BIOMARKER set were labeled by a certified Neurodiagnostic Technologist with annotations reviewed by a Neurologist. The labeler was assigned to identify any seizures as well as the first instance and peak sustained rate of the following features:

- Sporadic interictal epileptiform discharges (SIED)
- Lateralized periodic discharges (LPD)
- Generalized periodic discharges (GPD)
- Sharp wave discharges (SWD)
- Generalized paroxysmal fast activity (GPFA)
- Other rhythmic or periodic patterns.

Any features not observed were indicated as having a peak sustained rate of 0 events/minute.

### Genetics

To examine whether variant location relative to the GAP domain was associated with quantitative sleep and spectral EEG outcomes, participants with confirmed SYNGAP1 variants were classified into two groups based on exon position: pre/within-GAP (exons 1–15, encompassing the PH, C2, and RasGAP domains) and post-GAP (exons 16–19, encompassing the disordered regulatory region and coiled-coil domain). Variants within exons 1–15 were grouped together on the basis that this region encodes the structured PH, C2, and RasGAP domains, which function as an integrated catalytic unit. Variants of any type in this region — truncating, missense, or in-frame deletion — would be expected to disrupt catalytic GAP activity, either by removing downstream sequence or by destabilizing the domain structure required for function. In contrast, exons 16–19 encode a disordered regulatory region and coiled-coil domain. Variants there are expected to primarily disrupt protein-protein interactions and scaffolding rather than catalytic activity, regardless of variant type. Variant effect types (frameshift/splice, nonsense/stop, missense, in-frame deletion) were assigned based on reported variant consequence, with three ambiguous cases (SYNGAP-002, -017, -021) manually reviewed and classified. One participant with a whole-gene chromosomal deletion (SYNGAP-009) was excluded from domain-level analyses. Available sample sizes differed slightly across outcome domains due to data availability: sleep macroarchitecture analyses included 14 pre/within-GAP and 3 post-GAP participants, while spectral analyses included 15 pre/within-GAP and 4 post-GAP participants.

Between-group comparisons of the pre-/within-and post-GAP subgroups were tested per feature with Mann-Whitney U tests. Comparisons between the pre-/within-GAP, post-GAP, and control groups used pairwise Mann-Whitney U tests. Group central tendency was summarized as median (IQR), and effect sizes were reported as rank-biserial correlations. To evaluate whether sleep architecture and spectral activity differed between the genetic subgroups while accounting for age, linear mixed models were fit for each feature, with genetic subgroup, mean-centered age, and their interaction as fixed effects and a per-participant random intercept. An additional analysis compared device (Waveband) usability metrics between subgroups using Mann-Whitney U tests. Significance was reported both uncorrected as well as after applying Benjamini-Hochberg FDR correction for multiple comparisons across all analyses.

## Data Availability

The data that was collected and analyzed for this study are not publicly available due to patient privacy as well as contractual restrictions.

## Supplemental Figures

**Supplemental Figure 1.**
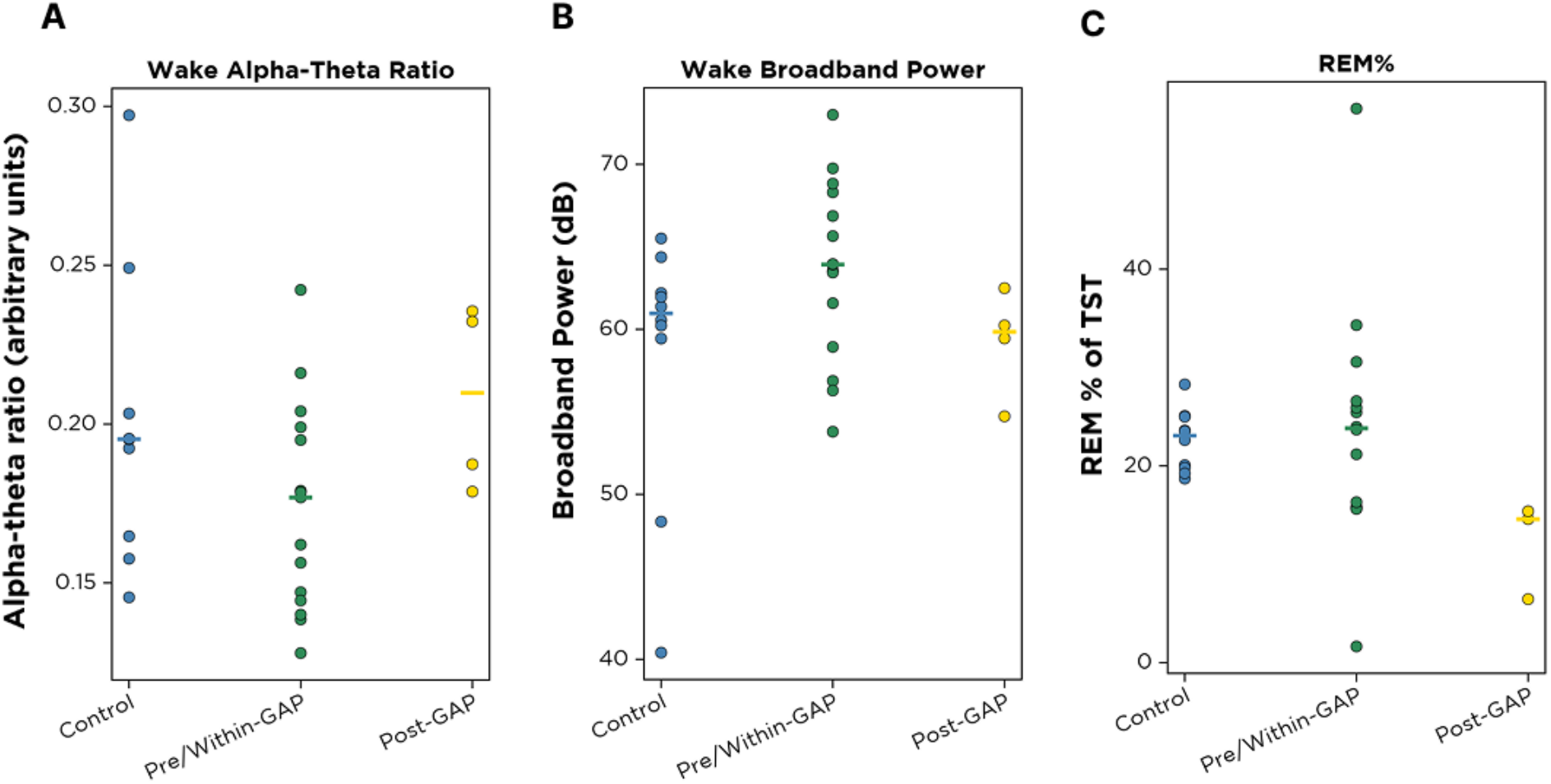
Individual-participant values for select spectral and sleep features, broken down into controls and genetic subgroups. Datapoints are individual SYNGAP1 participants grouped by GAP cluster and control participants (Control in blue, Pre/Within-GAP in green, Post-GAP in yellow); horizontal bars mark the group median. Given the small per-group sample sizes, panels show individual data points for (A) wake alpha-theta ratio, (B) wake broadband power, and (C) percentage of REM, to assess whether apparent group differences are driven by individual outliers.

